# EHRtemporalVariability: delineating temporal dataset shifts in electronic health records

**DOI:** 10.1101/2020.04.07.20056564

**Authors:** Carlos Sáez, Alba Gutiérrez-Sacristán, Isaac Kohane, Juan M García-Gómez, Paul Avillach

**Author notes:** Both to be regarded as last authors (Juan M García-Gómez, Paul Avillach).

## Abstract

**Background:** Temporal variability in healthcare processes or protocols is intrinsic to medicine. Such variability can potentially introduce dataset shifts, a data quality issue when reusing electronic health records (EHRs) for secondary purposes. Temporal dataset shifts can present as trends, abrupt or seasonal changes in the statistical distributions of data over time, being particularly complex to address in multi-modal and highly coded data. These changes, if not delineated, can harm population and data-driven research, such as machine learning. Given that biomedical research repositories are increasingly being populated with large historical data from EHRs, there is a need for specific software methods to help delineate temporal dataset shifts to ensure reliable data reuse.

**Findings:** EHRtemporalVariability is an Open Source R-package and Shiny-app designed to explore and identify temporal dataset shifts. EHRtemporalVariability estimates the statistical distributions of coded and numerical data over time, projects their temporal-evolution through non-parametric Information Geometric Temporal plots, and enables the exploration of changes in variables through Data Temporal Heatmaps. We demonstrate the capability of EHRtemporalVariability to delineate dataset shifts in three impact case studies, one of them available for reproducibility.

**Conclusions:** EHRtemporalVariability enables the exploration and identification of dataset shifts, contributing to broadly examine and repurpose large, longitudinal datasets. Our goal is to help ensure reliable data reuse to a wide range of biomedical data users. EHRtemporalVariability is suited to technical users programmatically using the R-package and to those users not familiar with programming using the Shiny user interface.

**Availability:** https://github.com/hms-dbmi/EHRtemporalVariability/ Reproducible vignette: https://cran.r-project.org/web/packages/EHRtemporalVariability/vignettes/EHRtemporalVariability.html On-line demo: http://ehrtemporalvariability.upv.es/

## Background

The widespread adoption of data-sharing technologies, health information standards, and open-data initiatives are inspiring the creation of research data repositories with large-scale historical data from EHRs^1^ This represents a new class of longitudinal, real-world data, defined as large datasets collected over time from sources outside of clinical trials or specific research cohorts. Reuse of this data, ranging from clinical observations to molecular information, has begun to boost the efficacy and generalization of biomedical and clinical research. These efforts, however, are still in early stages.^2,3^

Most recently researchers from the machine-learning community have identified EHR data as one of the most important sources of labeled data with which diagnostic and prognostic models can by constructed.^4^ One of the major hurdles in reusing such EHR data, however, is its temporal variability. Indeed, clinical care processes and their local variations are permeated with a variety of batch effects and biases.^5–9^ This is similar to the situation in genomics and other “omics” research, where batch effects can be introduced by technical sources of variation that have been added to samples during acquisition handling.^15,16^

These artifacts—in form of dataset shifts—can impact data quality and challenge the secondary use of data, particularly for population and data-driven research,^8,10–12^ as well as machine-learning.^13,14^

In addition, when dealing with research data repositories populated by EHRs, there are other sources of temporal variability. The EHR’s themselves also contribute to variability, as they reflect the evolution of administrative practice and reimbursement policies, all of which can gradually or abruptly shift over time. For example, updates in coding systems, such as the International Classification of Diseases (ICD),^17^ or modifications to clinical guidelines often lead to variable data representations across multiple diseases over time.

To circumvent these issues, researchers have traditionally deployed Statistical Process Control-based methods, which expose the timepoints when reference changes had occurred. Shewhart and Levey-Jennings charts, for example, have been employed in laboratory quality control efforts. Similarly, autocorrelation or time-series-based approaches have been used to uncover periodicity and changes within summary statistics derived from longitudinal samples, such as batched averages. Indeed, when the dates of such reference changes are known, statistical tests can uncover significant differences between time periods. However, these approaches tend to promulgate loss of information, especially when deployed when using categorical variables with a particularly high number of values. This is true of EHR-coded clinical data, such as the ICD Ninth Revision, Clinical Modification (ICD-9-CM), which has over 16,000 distinct codes, and in multimodal statistical distributions, in which multiple sub-phenotypes are present.

## Methods

EHRtemporalVariability is designed to explore and identify the temporal variability of categorical and numerical data over time. The app provides the means to visually and analytically delineate dataset shifts in multi-modal and highly coded information. A key advantage is that no distributional assumptions are made. This enables straightforward use, as well as visual analytics on large EHR-coded and numerical variables with no loss of information. In addition, the tool’s methodological and iterative use can identify and define reference changes that might otherwise impede further research. This can be done without concern about complex variable processing.

EHRtemporalVariability is based on the probabilistic temporal variability methods that we developed and validated over 5 years,^6,9,11^ namely Information-Geometric-Temporal (IGT) plots and Data Temporal Heatmaps (DTHs). We offer these for the first time as an open-source R package and Shiny app. Our method is based upon the estimation and comparison of data statistical distributions over time (see online Methods). IGT plots project time batches as a series of points. The distances between them correspond to the dissimilarity of their statistical distributions. This yields an empirical layout of temporal relationships between batches, namely a non-parametric temporal statistical manifold.

IGT plots allow users to visually identify four types of changes: trends, represented as continuously flowing time batches; abrupt changes, shown as gaps between groups of batches; temporal subgroups, depicted as clusters of batches; and seasonality, portrayed as temporal cycles. Additionally, batches are labeled by date and color-coded to distinguish seasonal effects. The IGT plot data also provides the means to identify those changes in order to model seasonal effects or apply clustering methods to depict temporal subgroups.^9^ Complementing the IGT plots, DTHs allow users to explore changes in absolute and relative frequencies over time—and at multiple variable values, simultaneously (e.g., frequencies of phenotypes).

Overall, the EHRtemporalVariability R package (**Figure 1, left**) and Shiny app (**Figure 1, right**) provide a set of functionalities that allow users to perform three actions: loading and processing datasets; running batched data analyses for estimation of DTHs and IGT projections; and visualizing these data through interactive plots. The R package also enables users to conduct these tasks programmatically, enabling more flexibility in data processing and further analysis of the resultant objects and embedding matrices.

**Figure 1.**
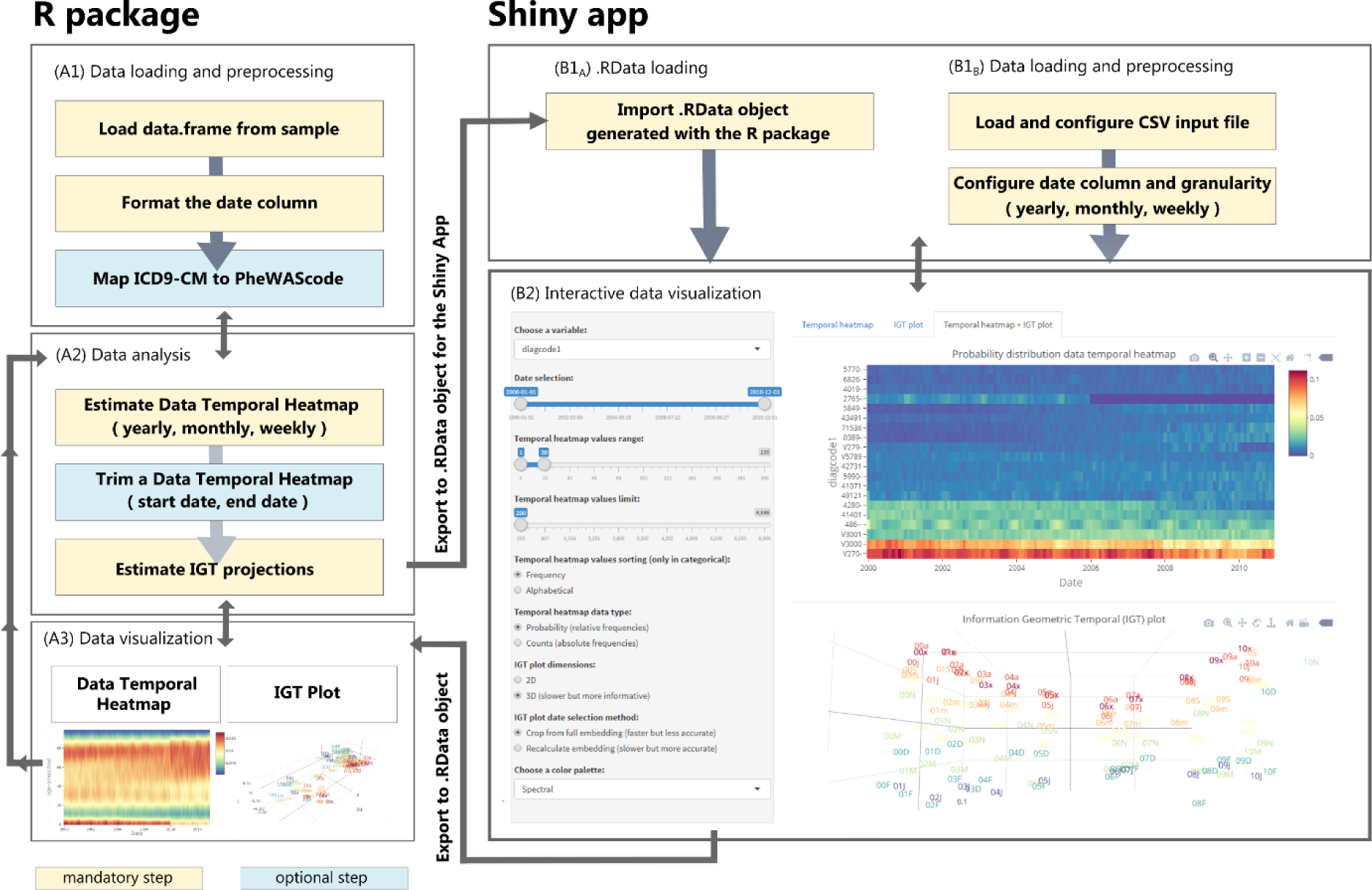
EHRtemporalVariability R package (left) and Shiny app (right) outline. The general workflow of the R package is organized as a set of functions for: (A1) data loading and preprocessing, (A2) data analysis, and (A3) data visualization. The main input is an R *data.frame*, in which one column defines the reference date. The classes of the remaining columns determine the variable’s treatment for distribution estimation and plotting during analysis and visualization (See online Methods section). Specifically, “factor” and “character*”* receive categorical treatment, while “numeric,” “*i*nteger,*”* and “date” receive numerical treatment. The DTH object estimation takes the input “data.frame*”* and analysis parameters. These include temporal granularity; predefined distribution support (a range of possible values or bins for each variable, auto-calculated from data by default); handling of missing batches; and the choice of whether to smooth distributions in numerical variables. The DTH can be trimmed by values and date range. The IGT projection estimation takes as input the DTH and the desired number of dimensions for embedding. The DTH can be plotted as a dynamic Plotly heatmap, in which the color of each cell indicates the frequency (relative or absolute) at a specific date batch (column) for the value of a variable (categorical and numerical integer) or range or values (numerical continuous). IGT plots can be visualized as either two or three-dimensional dynamic Plotly plots. The input for the Shiny app can be either an *.RData* object exported from the R package (B1A) or a raw .csv input file (B1B). The Shiny app provides an interactive dashboard (B2) for controlling the visualization parameters of the programmatic R functions. This is done via reactive sliders, selection boxes, and buttons. These have a direct effect on heatmaps and IGT plots. Further, we include different color palettes suited for different types of colorblindness. For further information about all the EHRtemporalVariability functionality see: https://cran.r-project.org/web/packages/EHRtemporalVariability/vignettes/EHRtemporalVariability.html.

The Shiny app provides a graphical user interface with two objectives. First, users unfamiliar with R programming can load *.csv* files and easily produce and visualize their results—which can be exported as a *.Rdata* file for further inspection in R. Second, we provide an exploratory, dynamic dashboard to improve the user experience, enabling a means to load results exported from the R package as a *.RData* file. We customized both the R package and Shiny app visualizations for users who are colorblind.

A more detailed description of methods is available in the Supplementary Material of the paper.

## Results and discussion

We validated the functionality of EHRtemporalVariability via three case studies. First, the i2b2 Boston Children’s Hospital Autism Spectrum Disorders cohort (BCH-ASD), including 12,000 patients (1.2M ICD-9-CM clinical observations), recorded from 1981 to 2016. This project was reviewed by Boston Children’s Institutional Review Board.

In this cohort, the IGT plot uncovered five changes of reference (**Figure 2a**). The most obvious was in billing codes, for which frequencies changed in October 1998 (**Figure 2a-a2**). Accordingly, we discovered an abrupt change in the relative frequencies of ICD-9-CM codes during that month. Specifically, the DTH of the ICD-9-CM codes (Supplementary Material Fig 1) showed an abrupt decrease in the frequencies of codes: 780 (general symptoms), 780.9 (other general symptoms) and 289.9 (unspecified diseases of blood and blood-forming organs). We also tracked increases in more specific 780.x codes; 296.x codes (episodic mood disorders), and other gradual changes.

**Figure 2.**
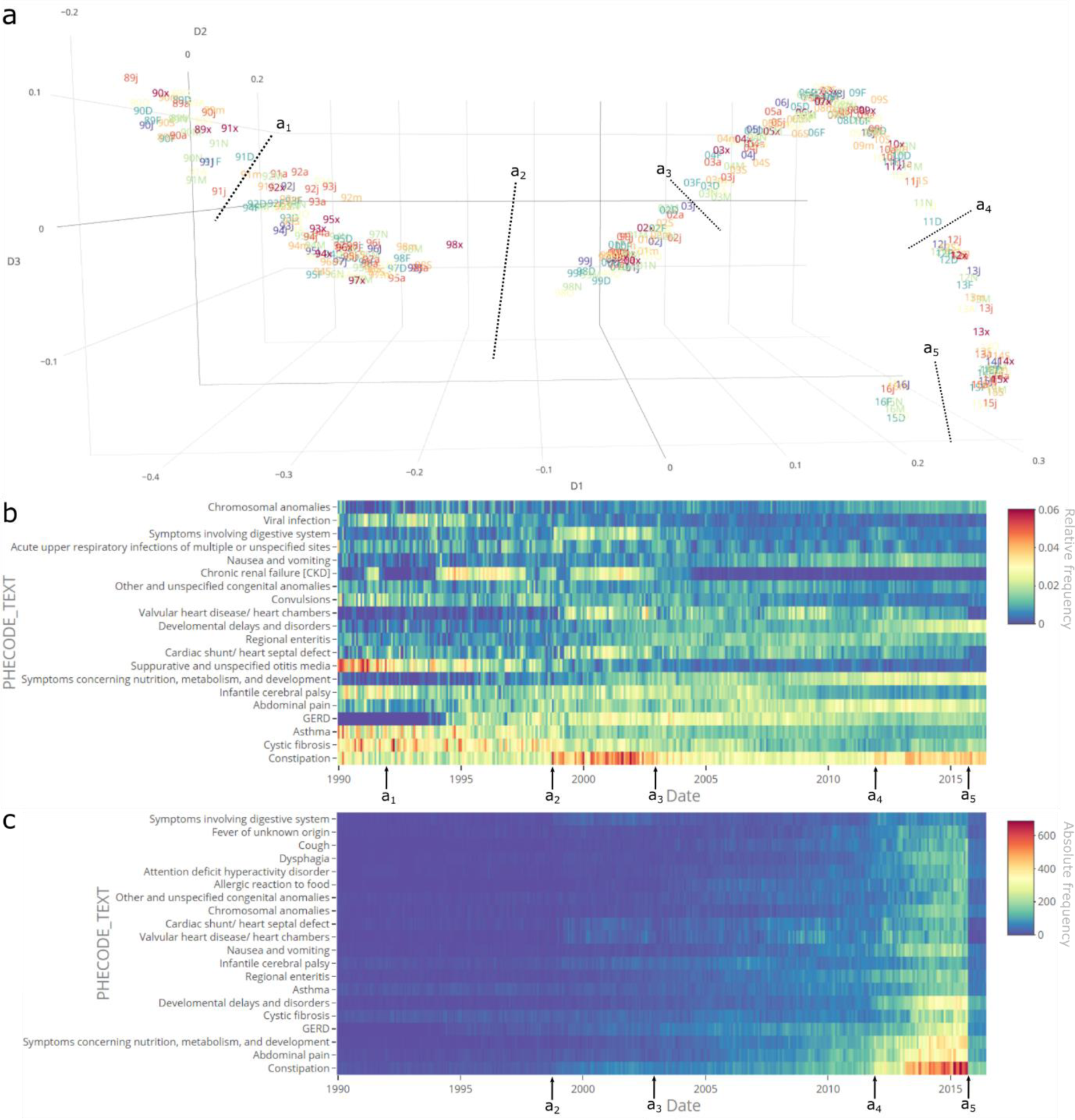
Delineation of dataset shifts in the Boston Children’s Hospital EHR Autism Spectrum Disorders historical clinical observations. (**a**) IGT plot describing the evolution of distributions of ICD-9-CM codes over time—specifically monthly time batches, taken from March 1989 to June 2016. The projection of time batches is based on embedding the dissimilarities among their distributions using multi-dimensional scaling. The IGT plot axis corresponds to the three first temporal components of variance. Several slight changes of reference are apparent during October 1991 (a_1_), January 2003 (a_3_), and December 2011 (a_4_). Major changes of reference appear during October 1998 (a_2_) and October 2015 (a_5_). Overall, there is a trend in the distribution changes across all the entire time frame. Text labels are formatted as “yym,” where “yy” is a two-digit year and “m” is an abbreviated month, as {‘J’, ‘F’, ‘M’, ‘A’, ‘m’, ‘j’, ‘x’, ‘a’, ‘S’, ‘O’, ‘N’, ‘D’}. (**b**) Data Temporal Heatmap of the 20 most frequent relative frequencies of PheWAS codes text. (**c**) Data Temporal Heatmap of the 20 most frequent absolute frequencies of PheCodes text. The major driver for (a_2_) was a decrease in “other symptoms” and “other tests” codes. Thus, we excluded these to investigate the effect on comorbidities and obtained (b) and (c). Changes in October 1998 (a_2_) include increases in the frequencies of “constipation,” “major depressive disorder,” “symptoms involving digestive system,” and “type 2 diabetes.” Other minor decreases included “cystic fibrosis,” “other diseases of blood and blood forming-organs,” and “type 1 diabetes,” among “others. As observed in (c), some of the delineated changes are time-correlated with alterations in absolute frequencies.

While investigating the root cause of the October 1998 reference change, we found that it coincided with a yearly ICD-9-CM update. However, there was no apparent relationship between documented changes and our findings.

To further investigate this variability, we mapped ICD-9-CM to Phenome Wide Association Studies (PheWAS) codes.^18^ We removed all the observations listed as “other symptoms” and “other tests.” Still, the abrupt change persisted even when delineated changes for further specific comorbidities (**Figures 2b, 2c**). Intriguingly, the absolute number of observations also increased at the start of the month. Although this reference change appears to be motivated by a systemic or protocol change, the exact cause remains unclear. We suggest that this reference change is a potential dataset shift that should be considered in any future BCH-ASD data analysis.

The second case study replicates a baseline experiment we previously performed using the Mortality Registry of Valencia, Spain.^11^ The registry recorded 512,000 deaths between 2000 and 2012. Similar to the Boston Children’s results, the registry’s statistical distributions changed abruptly in 2009, following a change in the fields of the Spanish National Death Certificate. Notably, this reference change impacted the Basic Cause of Death, a variable used for reporting National and International death statistics (**Figure 3** and Supplementary Material Fig 2). This occurred even after the variable was retrospectively corrected.

**Figure 3.**
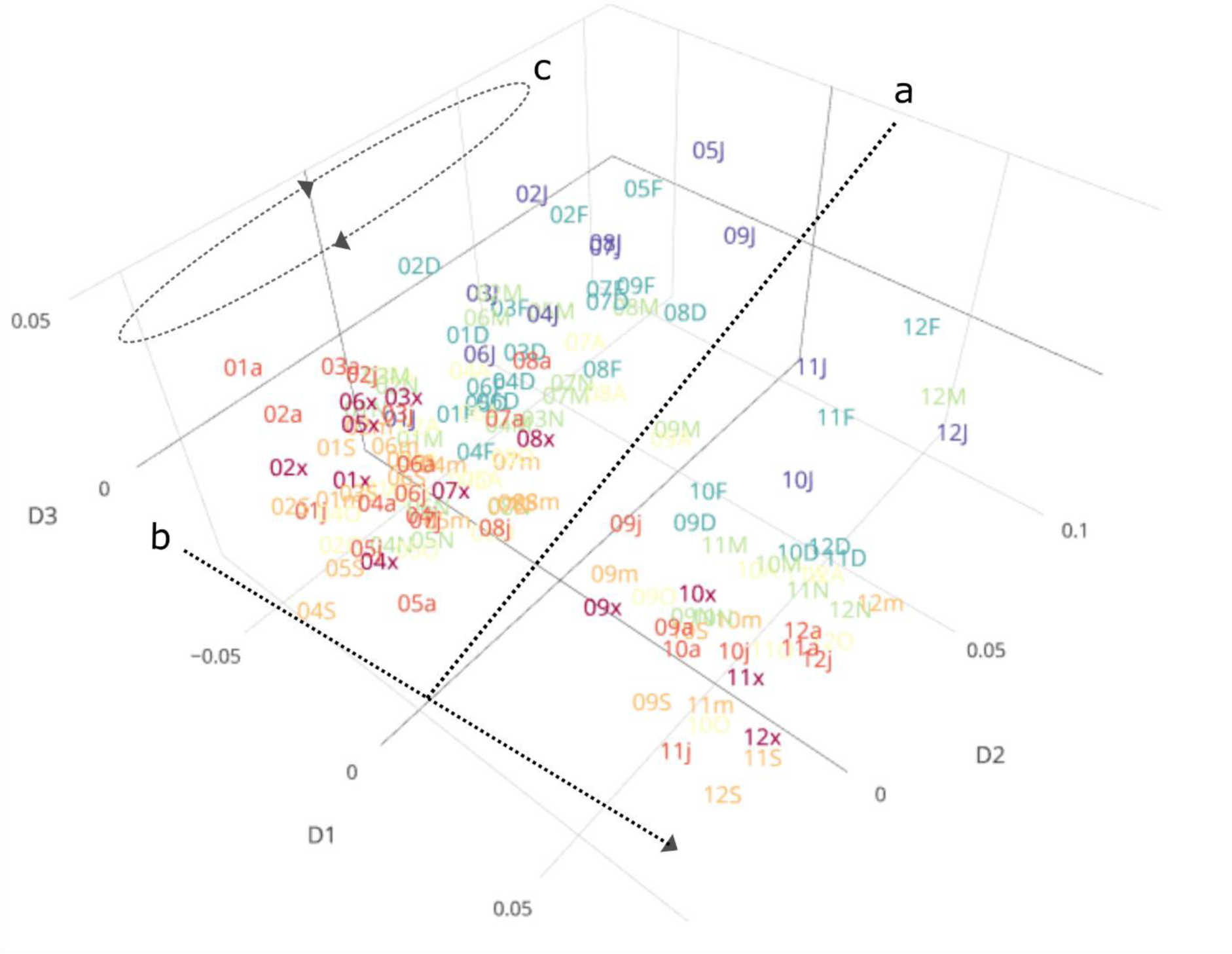
IGT plot of the Basic Cause of Death in the Mortality Registry of the Region of Valencia, Spain, coded with ICD Tenth Revision (ICD-10) Mortality Causes List 1. (**a**) The major abrupt change associated with the update of the National Certificate of Death is depicted as a dotted line that splits the main trend through the entire period of study, (**b**) layering on dimension D1. (**c**) Yearly seasonality of causes of death, highlighted by coloring scheme and laid out across dimension D2. Text labels are formatted as”‘yym,” where “yy” is a two-digit year and “m” is an abbreviated month as {‘J’, ‘F’, ‘M’, ‘A’, ‘m’, ‘j’, ‘x’, ‘a’, ‘S’, ‘O’, ‘N’, ‘D’}. The drivers for (a) included a relatively abrupt decrease in “symptoms, signs, and abnormal clinical and laboratory findings, not elsewhere classified” and an increase in “hypertensive diseases,” among others.

Finally, we validated EHRtemporalVariability with the National Hospital Discharge Survey (NHDS), an open dataset that includes 3.25M inpatient discharges from US hospitals (2000-2010) and both demographic and ICD-9-CM-coded data. Again,^6,9^ we uncovered several abrupt reference changes throughout multiple variables, including the re-coding of discharge age in 2008; ICD-9-CM diagnosis (**Figure 1**); procedure codes; and yearly abrupt changes in diagnosis-related group codes. These findings were in addition to the expected context-induced trends and seasonality. After mapping the NHDS ICD-9-CM codes to PheWAS codes, we noted that notable changes remained, including those appearing in October 2007, coincident with the yearly ICD-9-CM update. Note: this case study is available for replication within the package and Shiny app demonstration at http://ehrtemporalvariability.upv.es/. Performance measures for the three case studies are described in the Supplementary Material.

## Conclusions

In conclusion, EHRtemporalVariability is a data quality assessment tool that provides the means to broadly explore and repurpose large data sets collected over time, and a stepping stone in the identification of dataset shifts for data reuse, specially in machine-learning. Target users are biomedical data scientists and bioinformaticians, as well as epidemiologists and hospital data managers. The tool can assist in exploring the effects of system, protocol, and environment-induced changes on data. We also encourage the use of EHRtemporalVariability to analyze the impact of the adoption of new coding systems such as ICD-10.^19^ EHRtemporalVariability can be used on any additional coded and numerical data modalities and, because it is open-source, the app can be extended with new functionality or uses by the scientific community.

## Data Availability

The data of the NHDS case study is publicly available at https://www.cdc.gov/nchs/nhds/index.htm. A random subset of this dataset is available as a proxy for testing purposes within the EHRtemporalVariability package, and reproducible examples are available within the package help, its vignette, and the on-line demo. Access to BCH-ASD case study data is restricted by Boston’s Children’s Institutional Review Board. Access to the Mortality case study data is restricted by the Conselleria de Sanitat Universal i Salut Pública, Generalitat Valenciana, Spain.

## Availability of code and resources

- Project name: EHRtemporalVariability
- Project home page: https://github.com/hms-dbmi/EHRtemporalVariability/
- Operating system(s): Platform independent
- Programming language: R
- Other requirements: R 3.3.0, dplyr, plotly, shiny, zoo, xts, lubridate, RColorBrewer, viridis, scales, methods
- License: Apache License 2.0
- CRAN repository: https://cran.r-project.org/package=EHRtemporalVariability
- Shiny-app repository: https://github.com/hms-dbmi/EHRtemporalVariability-shiny
- Reproducible vignette: https://cran.r-project.org/web/packages/EHRtemporalVariability/vignettes/EHRtemporalVariability.html
- On-line Shiny app demo (for privacy reasons loading raw .csv data is disabled): http://ehrtemporalvariability.upv.es/

## Additional files

We provide a Supplementary material file including (1) the technical details of the methods, (2) supplementary figures and (3) a performance measures test.

## Abbreviations

BCH-ASD: Boston Children’s Hospital Autism Spectrum Disorders cohort
DTH: Data Temporal Heatmap
EHR: Electronic Health Record
ICD: International Classification of Diseases
ICD-9-CM: ICD Ninth Revision, Clinical Modification
IGT plot: Information Geometric Temporal plot
NHDS: National Hospital Discharge Survey
PheWAS: Phenome Wide Association Studies

## Acknowledgments

This work was supported by UPV grant PAID-00-17, GVA grant BEST/2018, and projects H2020-SC1-2016-CNECT No. 727560 and H2020-SC1-BHC-2018-2020 No. 825750. The authors thank the community that collaboratively created the Open Source R software and packages used in this work. A special thanks to UpSetR, which inspired our Shiny wrapper landing page.

